# Effect of Tai Chi on Pain and Cognitive Function in Elderly Patients with Chronic Neck Pain: A Protocol for Systematic Review and Meta-analysis

**DOI:** 10.1101/2023.07.10.23292456

**Authors:** Yichen Wang, Xing Tang, Hongjuan Fu, Wenjiao Hu, Feng Zhang

## Abstract

**Background:** The incidence of chronic neck pain (CNP) in the elderly is increasing. CNP in the elderly not only affects the physical function of patients, the quality of life, and work but also increases the direct medical cost and indirect social costs. The recent treatment trends of CNP, such as home self-exercise, focused specific training, and research into the underlying neural mechanisms of pain and cognitive improvement in patients with CNP, suggest that Tai Chi may be beneficial for pain and cognitive function in elderly patients with CNP.

**Method and Methods:** We will search 8 databases, including PubMed, Embase, Cochrane Library, Web of Science, China National Knowledge Infrastructure (CNKI), China Biomedical Literature Database (CBM), VIP database, and Wanfang database. Randomized controlled trials (RCTs) of Tai Chi in the treatment of elderly CNP will be included. The search time is from the establishment of the database to May 31, 2023. In this study, data extraction and methodological quality evaluation will be carried out for the included literature, and statistical analysis will be performed using RevMan 5.4 software. The quality of evidence will be assessed using the Grading of Recommendations Assessment, Development and Evaluation system approach(GRADE).

**Discussion:** This review will search and collect RCTs related to Tai Chi for the prevention and treatment of elderly CNP, and aims to systematically evaluate the evidence of the effects of Tai Chi on pain and cognitive function in elderly patients with CNP. The results of this review are expected to provide more accurate and effective guidance for the prevention and treatment of elderly patients with CNP, and also provide evidence-based medicine references for the formulation of exercise prescriptions.

**Systematic Review Registration:** OSF registration number [DOI 10.17605/OSF.IO/YTZPX].

## Introduction

Chronic neck pain (CNP) in the elderly is a clinical syndrome mainly manifested as neck pain, stiffness, discomfort, and limited movement, accompanied by fatigue and discomfort, shoulder pain, head lethargy, and other symptoms, with a course of more than 12 weeks [1]. In 2010, among the global studies on life lost due to diseases, the number of years of life lost due to disability caused by neck pain ranked fourth [2].

Epidemiological research on CNP found that the incidence of CNP in the general population ranged from 0.055/1000 to 213/1000, with a high incidence of CNP between 40 and 60 years old, and the prevalence of CNP in older people over 70 years old could reach 90%[3]. A large-scale study on the prevalence and risk factors of CNP in the elderly in South Korea showed that the lifetime prevalence of CNP in the elderly in South Korea was up to 20.8%, and the prevalence was higher in females [4]. At present, the incidence of CNP in the elderly population remains high and increases year by year, which has become a social and medical problem in various countries.

There are many causes of CNP in the elderly, among which an important predictor is injury [5]. Common neck injuries from car accidents or falls from height. According to the literature, 20%-40% of whiplash neck injuries will gradually develop into CNP [6]. In addition, abnormal nerve control in the neck, muscle fatigue, skeletal muscle diseases caused by neck imbalance, improper posture, obesity, and mental stress are high-risk factors for CNP in the elderly[7]. Elderly people with CNP usually suffer from memory decline, attention loss, reaction ability slowed down and other normal physiological function decline, seriously affecting the patient’s physical function, life, and work quality[8].

Although oral medications, physical therapy, acupuncture, and manipulation can alleviate the symptoms of CNP in the elderly to varying degrees and improve the function of the cervical spine, it is difficult to fundamentally curb the development of CNP, and patients often need repeated treatment or even lifelong treatment. Data show that the prevalence rate of neck pain in people’s lifetime is about 40%-70%[9,10]. 33% to 65% of patients recover from neck pain within one year of onset, but recurrence is common [11]. Elderly patients with CNP often suffer from severe adverse emotions and psychological reactions due to recurrent and persistent neck pain [12], and gradually develop into emotional disorders with anxiety as the prominent manifestation, which will also affect patients’ confidence and efficacy in treatment [13]. It is of great social, economic, and medical significance to explore effective methods to treat and prevent CNP in the elderly.

There have been some recent advances in the treatment of CNP in the elderly. Lauche et al. [14,15] reported a randomized controlled trial (RCT) of Tai Chi in treating chronic non-specific neck pain, and the results showed that compared with the waiting treatment group, the Tai Chi training group could reduce the degree of neck pain and improve the dysfunction index. Two expert opinion literature [16,17] also stated that tai chi exercise can not only reduce the neck pain of patients, improve the function of patients, but also reduce the depression and anxiety caused by pain, which is very beneficial to the prevention and treatment of CNP. At present, Tai Chi has been applied in the treatment of various chronic diseases, and has been proven to have positive therapeutic effects [18].

There is growing evidence showing that tai chi also helps cognitive function in elderly patients with CNP, which may be partly attributable to remodeling central nervous system activity [19]. The elderly aged 60-70 years old were divided into a swimming group, Tai Chi group, square dancing group, and control group, and the results showed that Tai Chi had the most significant effect on the cognitive function of the elderly [20]. Long-term Tai Chi practitioners score higher on the cognitive scale, and longer exercise time can improve the cognitive function of the elderly [21,22].

Recent advances have also been made in understanding the pathophysiology of CNP in the elderly. Imaging studies have shown that the relationship between different types of chronic pain and brain regions corresponds to a complex correspondence. Patients with chronic pain have abnormal activation of multiple brain regions and functional connectivity changes [23,24]. Persistent chronic pain can significantly activate not only pain-inducing centers but also cognitive and emotional brain regions [25]. For example, studies on CNP found that connections in brain regions such as anterior cingulate gyrus, posterior cingulate gyrus, precuneus, parietal gyrus, inferior parietal gyrus, thalamus, bilateral insula, and frontal lobe were enhanced compared with healthy volunteers [26]. In addition, one study showed that the left precuneus cortex was thinner in traumatic CNP compared to non-traumatic patients, and patients with non-traumatic CNP performed poorly compared to healthy volunteer controls after a measure of cognitive performance [27].

At present, Tai Chi has been applied in the treatment of various chronic diseases, and has been proven to have positive therapeutic effects [28]. To date, there have been no systematic reviews focusing only on the effectiveness of Tai Chi for CNP in the elderly. Elderly CNP has the characteristics of high incidence, complex symptoms, lingering disease, and easy relapse. Exploring the effective treatment and prevention of CNP in the elderly has obvious social, economic, and medical significance, and tai chi on the elderly CNP is worth studying. The purpose of this systematic review will be to identify and evaluate the evidence on the effects of Tai Chi on pain and cognitive function in elderly patients with CNP.

## Methods

### Study registration

This systematic review protocol was registered on the OSF (https://doi.org/10.17605/OSF.IO/YTZPX). The systematic review is designed to be by the Preferred Reporting Items for Systematic Reviews and Meta-Analyses (PRISMA) guidelines [29].

### Inclusion criteria

All RCTs on CNP in the elderly will be included that have tai chi alone or as a component of an intervention and have a comparison group, such as no treatment, regular activity, aerobic exercise, western medicine treatment, and other treatments. All participants in such studies will be included regardless of sex, nationality, or whether they are inpatients or outpatients. All types of Tai Chi and lengths of intervention will be included (Table 1).

**TABLE 1.** Inclusion criteria.

|  |
| --- |
| **Participants** |
| • Elderly patients with a definite diagnosis of CNP, aged $\geq 60$ years old, regardless of gender, race, nationality, and duration of disease. |
| **Intervention** |
| • Interventions in the treatment group will include different types of frequency and duration of Tai Chi therapy. |
| **Comparators:** |
| • Ordinary exercise unrelated to Tai Chi or blank control, etc. |
| **Outcome measures :** |
| • Primary outcomes: VAS, NPQ, MPQ, MMSE; |
| • Secondary outcomes : Neck Disability Index (NDI) , the incidence of adverse events. |
| **Design** |
| • Randomized clinical trials |

### Outcome measures

As per recent trends to have patient-oriented outcomes, we have identified the preferred primary outcome as pain and cognition, such as Visual Analogue Score (VAS), Northwick Park Questionnaire (NPQ), Mcgill Pain Questionnaire (MPQ), Mini-Mental State Examination (MMSE). Secondary outcomes are neck function and any adverse events.

### Search strategy

We will search RCTs on the effects of Tai Chi on pain symptoms or cognitive function in elderly patients with CNP in PubMed, Web of Science, The Cochrane Library, Embase, CNKI, CBM, Wanfang, and VIP databases. The time limit is to build the database until May 31, 2023. The combination of subject words and free words is adopted. Detailed search strategies are shown in Table 2.

**TABLE 2.** Search strategy in PubMed database.

| Search items |  |
| --- | --- |
| 1 | Tai Ji. Mesh. |
| 2 | Tai Chi. ti. ab. |
| 3 | Chi, Tai. ti. ab. |
| 4 | Tai Ji Quan. ti. ab. |
| 5 | Ji Quan, Tai. ti. ab. |
| 6 | Quan, Tai Ji. ti. ab. |
| 7 | Taiji. ti. ab. |
| 8 | tai chi. ti. ab. |
| 9 | Tai Chi Chuan. ti. ab. |
| 10 | 1 or 2–9 |
| 11 | Aged. Mesh. |
| 12 | Elderly. ti. ab. |
| 13 | 11 or 12 |
| 14 | Neck Pains. Mesh. |
| 15 | Pain, Neck. ti. ab. |
| 16 | Pains, Neck. ti. ab. |
| 17 | Neck Ache. ti. ab. |
| 18 | Ache, Neck. ti. ab. |
| 19 | Aches, Neck. ti. ab. |
| 20 | Neck Aches. ti. ab. |
| 21 | Cervicalgia. ti. ab. |
| 22 | Cervicalgias. ti. ab. |
| 23 | Cervicodynia. ti. ab. |
| 24 | Cervicodynias. ti. ab. |
| 25 | Neckache. ti. ab. |
| 26 | Neckaches. ti. ab. |
| 27 | Cervical Pain. ti. ab. |
| 28 | Cervical Pains. ti. ab. |
| 29 | Pain, Cervical. ti. ab. |
| 30 | Pains, Cervical. ti. ab. |
| 31 | 14 or 15–30 |
| 32 | Randomized controlled trial. Mesh. |
| 33 | Controlled clinical trial. ti. ab. |
| 34 | Randomized. ti. ab. |
| 35 | Randomly. ti. ab. |
| 36 | Trial. ti. ab. |
| 37 | 32 or 33–35 |
| 38 | 10 and 13 and 31 and 37 |

### Data collection and analysis

The literature retrieved from each database will be imported and downloaded into Endnote to automatically eliminate duplicate literature. The two researchers (YW and XT) will independently screen references. By reading the full text, the literature that do not meet the inclusion criteria was removed. For the literature with multiple drafts of the same experimental data, the initial experimental data or the updated experimental data will be retained, and other literature will be excluded; For incomplete or unextractable experimental data in the included literature, contact the original author by E-mail or telephone to obtain complete data. If the original author cannot be contacted or the original author cannot provide the required information and data, the literature will be excluded. If no agreement can be reached, it shall be settled through discussion and negotiation. If no agreement can be reached, it shall be settled by a third reviewer (FZ). The process of literature screening is shown in Figure 1.

**Figure 1.**
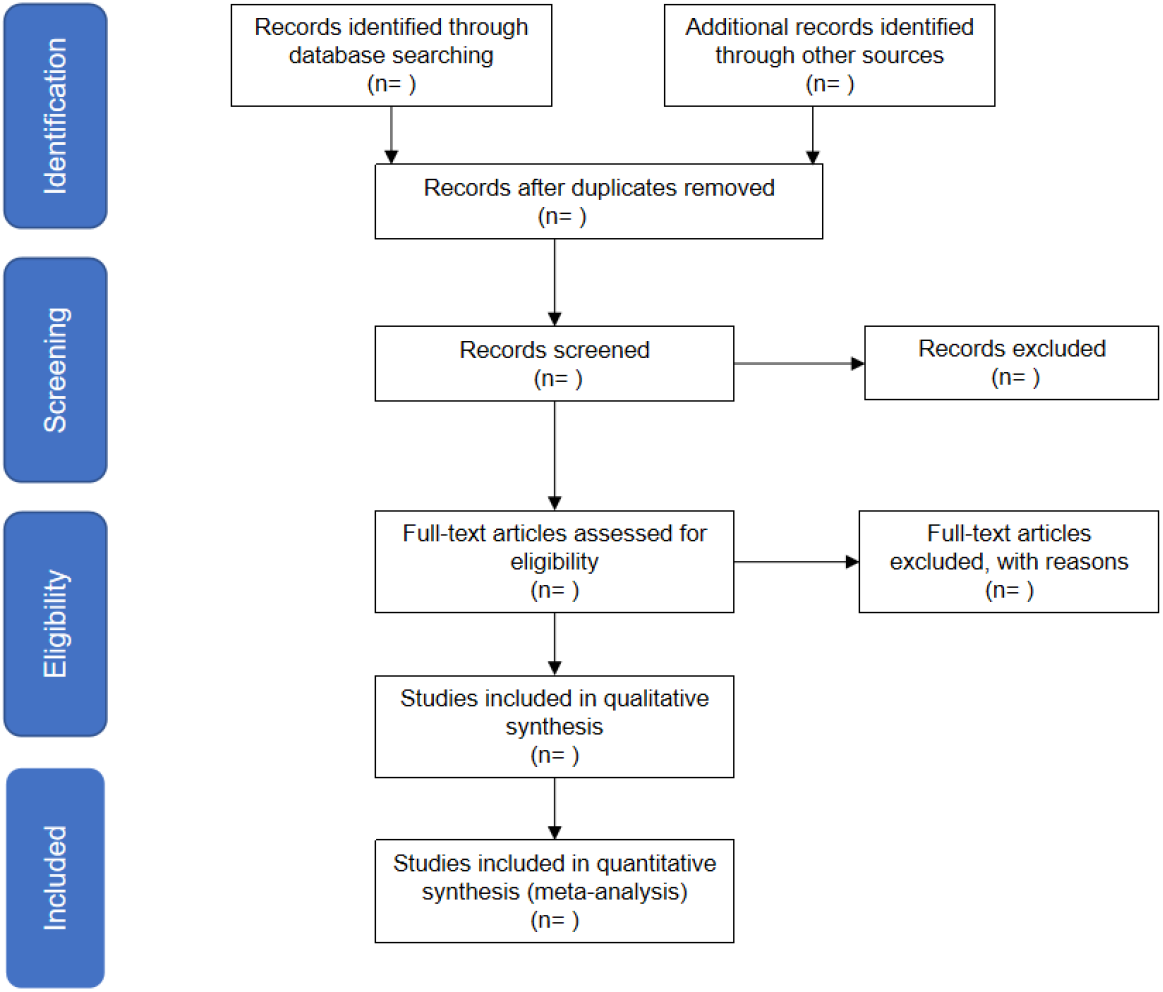
Flow diagram of the study selection process.

### Data extraction

Data will be extracted and cross-checked independently by two researchers (HF and WH) according to pre-established criteria. In case of disagreement, discuss or consult a third party (FZ). The content of data extraction included : (1) Basic information included in the study, namely the study title, the first author, the source of the literature, the publication time, and the intervention measures of the trial group and the control group ;(2) Baseline characteristics of the subjects (sample size, age, etc.) ; (3) Key elements of quality evaluation ; (4) Outcome indicators concerned and analysis results of each outcome indicator.

### Assessment of risk of bias

Two reviewers (YW and XT) will use the Cochrane risk bias assessment tool to evaluate the methodological quality of the included literature in six domains, such as selection bias, implementation bias, follow-up bias, measurement bias, reporting bias, and other biases. For each indicator, “low bias risk”, “bias uncertainty” and “high bias risk” were used to determine. The research quality was divided into three levels: Level A: Low risk met 4 or more items, that is, low risk; Level B: Meet 2 or 3 items of low risk; Level C: Meet 1 item or no entry low risk, deviation may occur. Any objection will be submitted to arbitration by the third reviewer (FZ).

### Assessment of heterogeneity

Data from the included literature will be analyzed using Revman5.4 statistical software available on the Cochrane Collaboration network. If the data are dichotomous variables, relative risk (RR) and 95%CI will be used as combined effect size analysis. If the data are classified as continuous variables, the mean difference (MD) or standard mean difference (SMD) and its 95%CI analysis were used. The chi-square test will be used to test the heterogeneity of the literature. When the heterogeneity of the included literature (I^2^ ≤ 50%) was determined, the fixed-effect model will be used to analyze the heterogeneity of the literature. When the heterogeneity of the included references will be greater than 50% (I^2^>50%), the random effects model will be used for analysis; if there are incomplete data, the descriptive analysis will be used.

### Assessment of reporting biases

According to the requirements of the Cochrane Systematic Review manual [30], funnel plots of Revman 5.4 software will be used to evaluate publication bias if ≥10 kinds of literature are included. If fewer than 10 articles are included, publication bias analysis will be not performed.

### Subgroup analysis and investigation of heterogeneity

We will use subgroup analysis to assess the heterogeneity of the results. Subgroup analysis will be performed by study design (such as intervention type, intervention duration, and intervention period), study location, and other factors to further reveal the impact of these factors on the study results or effect size.

### Sensitivity analysis

Sensitivity analysis will be used to evaluate the robustness of the meta-analysis conclusions. Each study will be excluded one by one to observe the changes in the results of the meta-analysis, and the results of other studies will be combined again with the effect size and compared with the previous total effect size to explore the robustness and influencing factors of the excluded studies on the results of the combined effect size. If there is no essential change in the meta-analysis results before and after the sensitivity analysis, it indicates that the meta-analysis results are more credible. If there is a large difference between the results before and after the sensitivity analysis, it indicates that the stability of the conclusions is poor, and the results and conclusions of the meta-analysis should be interpreted with caution.

## Discussion

To our knowledge, this will be the first systematic review of Tai Chi in the treatment of CNP in the elderly. In addition to the symptoms of neck pain, stiffness and discomfort, limited activity, and other symptoms, CNP in the elderly usually suffer from the decline of normal physiological functions such as memory decline, attention loss, slow reaction ability, and the incidence of the disease is high, the symptoms are complex, the disease is lingering and easy to relapse. At present, a drug intervention is often used for elderly patients with CNP in the clinic. However, the side effects of drug treatment are easy to damage the body’s organs. It has become an urgent need to seek a safe, economical, and green treatment method without side effects. Tai Chi is a traditional Chinese exercise, which has positive effects on pain symptoms and overall cognitive function of elderly patients with CNP, but its improvement effect is affected by age, gender, project characteristics, exercise cycle, exercise frequency, and exercise duration. In terms of the mechanism of action affecting the cognitive function of the elderly, although the research results are becoming more and more perfect and the influence mechanisms of different sports are similar, the influence of Tai Chi on the brain mechanism changes of pain and cognitive function still needs to be further studied.

In clinical randomized controlled trials, Tai Chi included different types of frequency and duration. The effects of different types of tai chi have not been synthesized [31]. Although a previous paper entitled “Tai Chi for Chronic Pain: A Systematic Review and Meta-analysis of Randomized Controlled Trials” was published [32], the disease was limited to chronic pain caused by osteoarthritis, rheumatoid arthritis, lumbago, fibromyalgia, and osteoporosis, and there has been no systematic review of Tai Chi for CNP in the elderly. In this systematic review, four English and four Chinese databases were searched, the risk of bias was assessed using the Cochrane bias risk tool, and GRADE was used to evaluate the evidence on the efficacy of Tai Chi in the treatment of CNP in the elderly. To provide relatively convincing evidence for the effectiveness and safety of Tai Chi in the treatment of CNP in the elderly. It can assist elderly patients with CNP and clinicians.

## Data Availability

No datasets were generated or analysed during the current study. All relevant data from this study will be made available upon study completion.

## Ethics and dissemination

There is no ethical conflict with the relevant content. The results of the meta-analysis will be published in a peer-reviewed journal.

## Author contributions

YW, XT and FZ conceived the review protocol. YW drafted the manuscript. YW and XT revised the study design. YW and XT will perform the study search and study selection. HF and WH will carry out the data collection. YW and XT will assess the quality of included randomised controlled trials. HF and WH will conduct the data analysis. FZ will monitor each procedure of the review and are responsible for the quality control. All authors have read and approved the publication of the protocol.

